# Companionship for women using English maternity services during COVID-19: National and organisational perspectives

**DOI:** 10.1101/2021.04.08.21254762

**Authors:** Gill Thomson, Marie-Clare Balaam, Rebecca Nowland, Nicola Crossland, Gill Moncrieff, Stephanie Heys, Arni Sarian, Joanne Cull, Soo Downe, ASPIRE-COVID19 Collaborative Group

## Abstract

**Objectives:** To explore the impact of COVID-19 on companionship for women using maternity services in England, as part of the Achieving Safe and Personalised maternity care In Response to Epidemics (ASPIRE COVID-19 UK) study.

**Setting:** Maternity care provision in England.

**Participants:** Interviews were held with 26 national governmental, professional, and service-user organisation leads including representatives from the Royal College of Midwives, NHS England, Birthrights and AIMS (July-Dec). Other data included public-facing outputs logged from 25 maternity Trusts (Sept/Oct) and data extracted from 78 documents from 8 key governmental, professional and service-user organisations that informed national maternity care guidance and policy (Feb-Dec).

**Results:** Six themes emerged: ‘*Postcode lottery of care*’ highlights variations in companionship practices, ‘*Confusion and stress around ‘rules’’* relates to a lack of and variable information concerning companionship, ‘*Unintended consequences’* concerns the negative impacts of restricted companionship on service-users and staff, ‘*Need for flexibility’* highlights concerns about applying companionship policies irrespective of need, ‘‘*Acceptable’ time for support’* highlights variations in when and if companionship was ‘allowed’ antenatally and intrapartum; and ‘*Loss of human rights for gain in infection control’* emphasizes how a predominant focus on infection control was at a cost to psychological safety and women’s human rights.

**Conclusions:** Policies concerning companionship have been inconsistently applied within English maternity services during the COVID-19 pandemic. In some cases, policies were not justified by the level of risk, and were applied indiscriminately regardless of need. This was associated with psychological harms for some women and staff. There is an urgent need to determine how to balance risks and benefits sensitively and flexibly and to optimise outcomes during the current and future crisis situations.

**Strengths and limitations of this study:**

- This is the first paper to consider links between policy and practice in companionship in maternity care during the COVID-19 pandemic;
- Data triangulation across stakeholders, policy and practice provides nuanced and context related perspectives on why and how companionship was impacted;
- Stakeholders included representatives from all key agencies involved in maternity care;
- Practice related issues were collected from the maternity Trust website and social media-based public facing information, which may or may not reflect actual care practices;
- The study does not include information directly reported by parents and healthcare professionals.

## INTRODUCTION

In many cultures around the world, pregnancy is framed as a social event rather than a clinical condition (1-3). Even where pregnancy and birth are framed as medically risky, social support is expected during pregnancy, birth, and the postnatal period. Such support is usually provided through ongoing family and community relationships, often by female relatives and friends, or community members (4). In the postpartum period most cultures recognise a period of between about 4 and 6 weeks during which the mother is supported to recover from the birth, to establish a relationship with the baby, and to adjust to motherhood (4).

As maternity care has become more hospital based, the power to determine who should accompany women in clinics and facility settings has shifted to the organisation, and its employees(5). In the early decades of mass hospitalisation for antenatal care and birth in the UK, companionship for women was banned or restricted, on the grounds of infection control, overcrowding, privacy for others, and defence from potential litigation if the accompanying partner or partners witness activities they perceive to be negligent or dangerous (6-8). More recently the evidence for the value of companionship in the intrapartum period has increased (1) and it is strongly recommended within global guidelines (9). Though restrictions still persist in some health economies around the world, antenatal, intrapartum and postnatal companionship policies in clinics and hospitals in the UK have become increasingly liberal over the last 40 years(10).

The COVID-19 pandemic brought the issue of companions in health facilities worldwide into sharp focus(11). In terms of maternity care, there have been anecdotal accounts of wide variations concerning if and whether women have been permitted companionship at various points throughout the maternity episode, both between countries, and across different care providers within countries. Concerns about women being alone for antenatal contacts, for ultrasound scans (especially when there is bad news), during labour and birth, and in the early postnatal period, have been widespread and global in media reports (for example: (11-13)).

To understand how companionship in maternity care during COVID-19 was operationalised organisationally in England, this paper presents an analysis of relevant national level policy documents; interviews with key stakeholders; and a review of public facing information produced by 25 purposively selected maternity providers in England.

## METHODS

The current study is part of a larger mixed-methods, observational, multi-site comparative study - Achieving Safe and Personalised maternity care In Response to Epidemics (ASPIRE COVID-19 UK).

Data relating to companionship in maternity care were extracted from national policy level documents and interviews with national stakeholders and mapped to analysis of public-facing communication channels from 25 Trusts (maternity care organisations). The Trusts were selected using maximum variation sampling, based on macro-level factors impacting upon health inequalities (area level deprivation): meso-level considerations relating to the organisation (Care Quality Commission (CQC) rating and maternal and neonatal mortality figures): and micro-level factors relating to the local population type and local organisational responses to the pandemic.

### Data collection

#### Documentary review

Guidelines, position papers, and reports relating to maternity care were collected prospectively between February and December 2020 from key governmental, professional, and service-user sources that inform national guidance and policy in relation to maternity care.

#### Trust-level public facing communication about maternity service provision

Data related to companionship in pregnancy, labour and birth, and the postpartum period were extracted from Trust websites and official Twitter, Facebook, and Instagram feeds, between September and October 2020.

#### Interviews

Individuals from relevant national governmental, professional, and service-user organisation leads involved in maternity care were approached by email and provided with an information sheet about the study. Semi-structured interviews were held July-December 2020 (using Microsoft Teams). Interviews lasted between 45-60 minutes, were audio recorded and transcribed in full. The interview schedule (see Supplementary File 1) explored stakeholders’ perceptions and experiences of what, why and how changes in maternity care delivery had been made during the pandemic, how changes had been monitored and assessed, and their views on facilitators and barriers to those changes. Interviews ceased when representation from all key organisations had been captured.

### Ethics approval

Ethics approval to undertake the interviews was obtained from the Health ethics sub-committee from the lead author’s institution (project no: 0079). Participants were asked if they would like to check any public facing quotes prior to publications as part of the consent process; quotes were returned to seven participants for feedback, and four required slight changes to be made.

### Data analysis

Data analysis was undertaken using a modified framework approach, using inductive and deductive methods (14). All the documents were searched for references to companionship. Relevant data segments were extracted and recorded in excel files. An initial data analysis framework was created by mapping the information from the national level documents into codes, and then collapsing them into key organising themes. This framework was then used to map and organise the interview data, with ongoing refinements to ensure that all the data were represented. The Trust-level data were subjected to descriptive content analysis (15). This involved the data being mapped to different aspects of care (e.g. appointments, ultrasound, induction, labour/birth). The Trust level data were then integrated into the thematic framework for reporting purposes. Four members of the team were involved in this process, and the final framework was agreed by all named authors.

### Public and Patient Involvement

This study was funded under a rapid response call and while no formal PPI involvement in the original design of the study, UK service-user leads (Maternity Voices Partnership) and members of third-sector organisations are involved as co-investigators, steering group and advisory members for the project, to ensure that service-user inputs has been considered at every stage of the study.

### Reflexivity

The authors and members of the collaborating group associated with this study are from a range of academic and clinical backgrounds including midwifery, psychology, obstetrics, neonatology, sociology, and social statistics. All the authors are female, and the four interviewers are experienced in undertaking qualitative interviews. One had previously collaborated with some of the interviewees. All authors believe that women and birthing people highly value companionship during key moments in their maternity care, and that, for many fathers/co-parents, being present is more than just being a visitor or a supporter. From her psychological background, RN also believes that birth supporters other than fathers/coparents play a significant role during childbirth in promoting psychological well-being. As midwives, SD, GM, JC, and SH view companionship for women throughout labour as a normative practice.

## RESULTS

A total of 50 individuals were approached to participate and interviews were held with twenty-six stakeholders (see Table 1). While some stakeholders did not respond to the requests, others provided names of individuals who they considered would be more suitable.

**Table 1.**
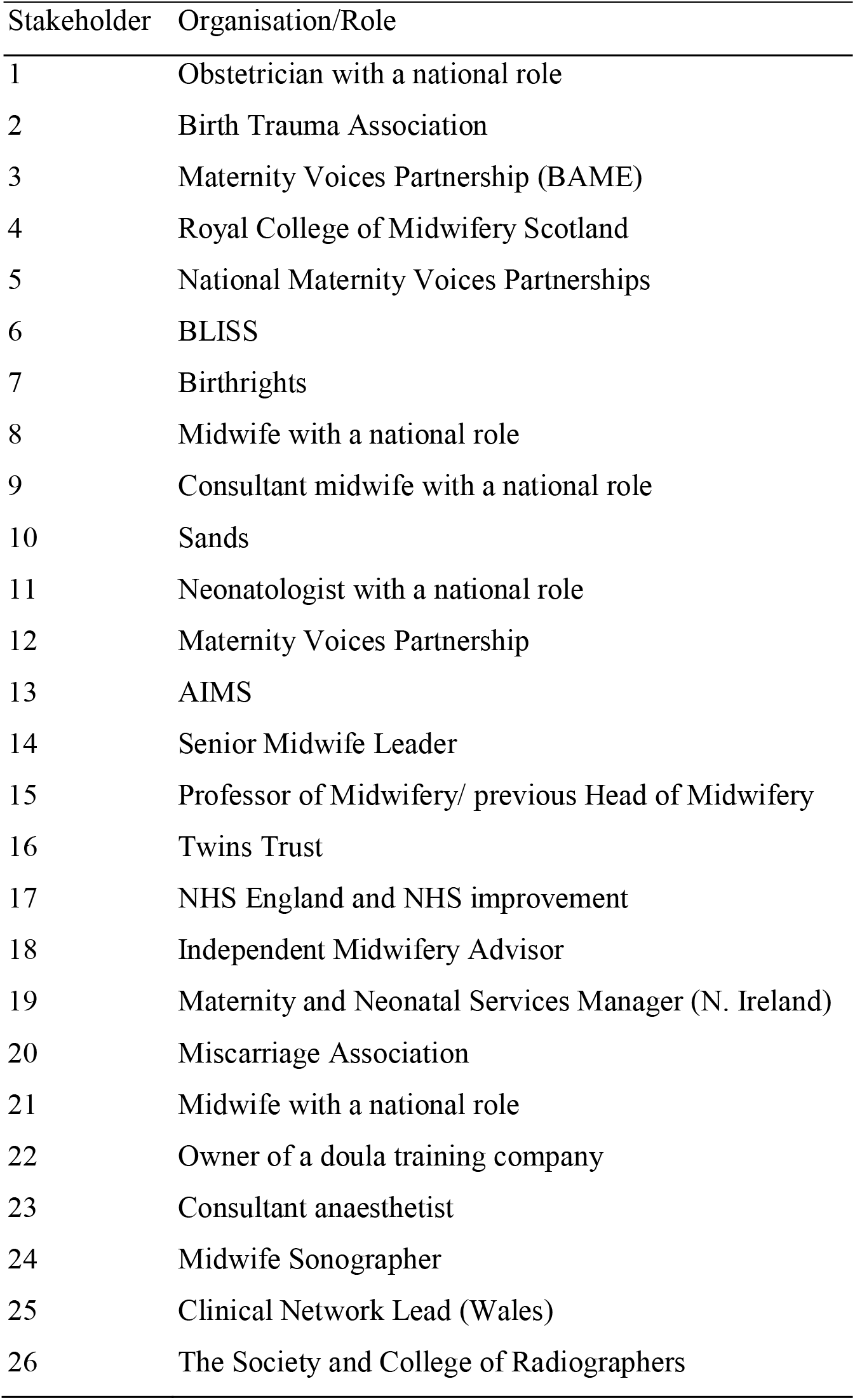
List of stakeholders interviewed

| Stakeholder | Organisation/Role |
| --- | --- |
| 1 | Obstetrician with a national role |
| 2 | Birth Trauma Association |
| 3 | Maternity Voices Partnership (BAME) |
| 4 | Royal College of Midwifery Scotland |
| 5 | National Maternity Voices Partnerships |
| 6 | BLISS |
| 7 | Birthrights |
| 8 | Midwife with a national role |
| 9 | Consultant midwife with a national role |
| 10 | Sands |
| 11 | Neonatologist with a national role |
| 12 | Maternity Voices Partnership |
| 13 | AIMS |
| 14 | Senior Midwife Leader |
| 15 | Professor of Midwifery/ previous Head of Midwifery |
| 16 | Twins Trust |
| 17 | NHS England and NHS improvement |
| 18 | Independent Midwifery Advisor |
| 19 | Maternity and Neonatal Services Manager (N. Ireland) |
| 20 | Miscarriage Association |
| 21 | Midwife with a national role |
| 22 | Owner of a doula training company |
| 23 | Consultant anaesthetist |
| 24 | Midwife Sonographer |
| 25 | Clinical Network Lead (Wales) |
| 26 | The Society and College of Radiographers |

Documents were collected from nine governmental, professional, and service-user sources (Box 1), with a total of 78 documents providing evidence for the paper (see Supplementary File 2 for full details/references).

### Box 1

Organisations included for documentary analysis

Sands

AIMS

Royal College of Midwives (RCM)

Royal College of Obstetricians and Gynaecologists (RCOG)

Society of Radiographers (SoR)

International Society of Ultrasound in Obstetrics and Gynaecology (ISUOG)

NHS England (NHSE)

Birthrights (BR)

The public facing data logged between September and October 2020 demonstrated a very wide range of policies and practices between the 25 included Trusts for companionship during four specific maternity care episodes (antenatal scanning; antenatal appointments; antenatal ward stays; and intrapartum) (see table two). While this could be explained by different COVID-19 infection exposure rates, this may not explain variation between Trusts in the same region.

**Table 2:**
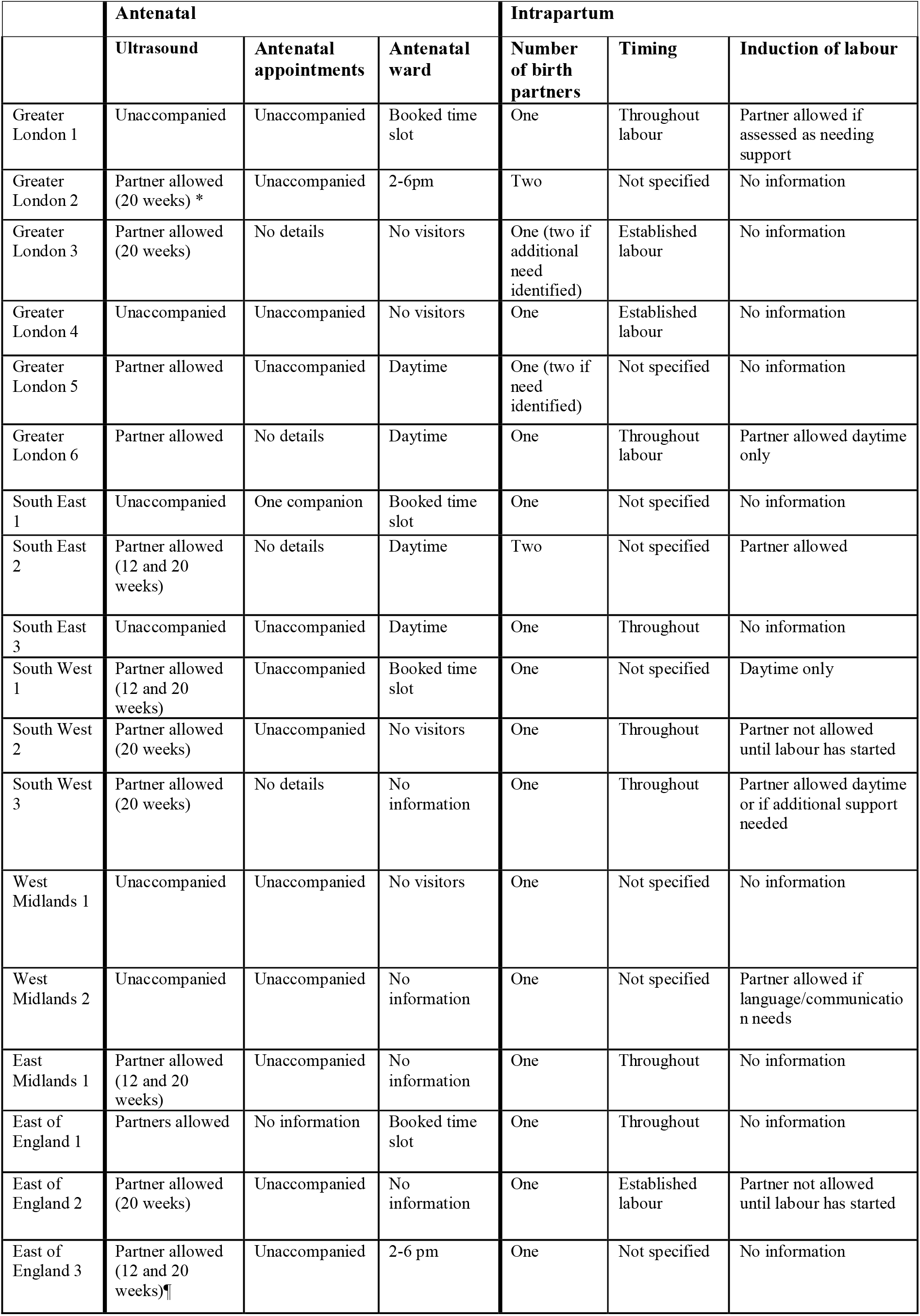
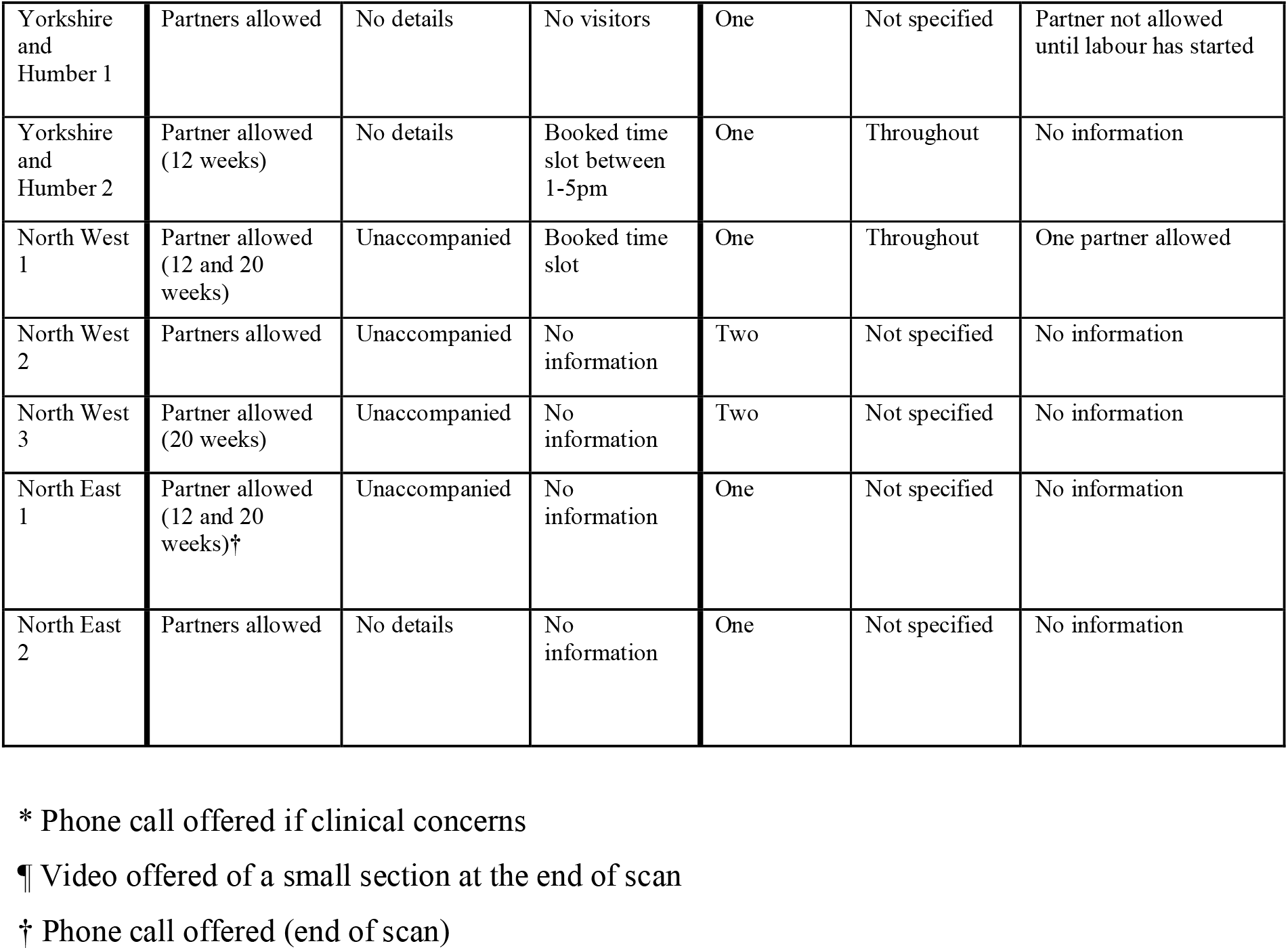
**Public facing information on maternity service companionship policies in 25 English maternity trusts, September to October 2020**

|  | **Antenatal** |  |  | **Intrapartum** |  |  |
| --- | --- | --- | --- | --- | --- | --- |
|  | **Ultrasound** | **Antenatal appointments** | **Antenatal ward** | **Number of birth partners** | **Timing** | **Induction of labour** |
| Greater London 1 | Unaccompanied | Unaccompanied | Booked time slot | One | Throughout labour | Partner allowed if assessed as needing support |
| Greater London 2 | Partner allowed (20 weeks) * | Unaccompanied | 2-6pm | Two | Not specified | No information |
| Greater London 3 | Partner allowed (20 weeks) | No details | No visitors | One (two if additional need identified) | Established labour | No information |
| Greater London 4 | Unaccompanied | Unaccompanied | No visitors | One | Established labour | No information |
| Greater London 5 | Partner allowed | Unaccompanied | Daytime | One (two if need identified) | Not specified | No information |
| Greater London 6 | Partner allowed | No details | Daytime | One | Throughout labour | Partner allowed daytime only |
| South East 1 | Unaccompanied | One companion | Booked time slot | One | Not specified | No information |
| South East 2 | Partner allowed (12 and 20 weeks) | No details | Daytime | Two | Not specified | Partner allowed |
| South East 3 | Unaccompanied | Unaccompanied | Daytime | One | Throughout | No information |
| South West 1 | Partner allowed (12 and 20 weeks) | Unaccompanied | Booked time slot | One | Not specified | Daytime only |
| South West 2 | Partner allowed (20 weeks) | Unaccompanied | No visitors | One | Throughout | Partner not allowed until labour has started |
| South West 3 | Partner allowed (20 weeks) | No details | No information | One | Throughout | Partner allowed daytime or if additional support needed |
| West Midlands 1 | Unaccompanied | Unaccompanied | No visitors | One | Not specified | No information |
| West Midlands 2 | Unaccompanied | Unaccompanied | No information | One | Not specified | Partner allowed if language/communication needs |
| East Midlands 1 | Partner allowed (12 and 20 weeks) | Unaccompanied | No information | One | Throughout | No information |
| East of England 1 | Partners allowed | No information | Booked time slot | One | Throughout | No information |
| East of England 2 | Partner allowed (20 weeks) | Unaccompanied | No information | One | Established labour | Partner not allowed until labour has started |
| East of England 3 | Partner allowed (12 and 20 weeks)¶ | Unaccompanied | 2-6 pm | One | Not specified | No information |
| Yorkshire and Humber 1 | Partners allowed | No details | No visitors | One | Not specified | Partner not allowed until labour has started |
| Yorkshire and Humber 2 | Partner allowed (12 weeks) | No details | Booked time slot between 1-5pm | One | Throughout | No information |
| North West 1 | Partner allowed (12 and 20 weeks) | Unaccompanied | Booked time slot | One | Throughout | One partner allowed |
| North West 2 | Partners allowed | Unaccompanied | No information | Two | Not specified | No information |
| North West 3 | Partner allowed (20 weeks) | Unaccompanied | No information | Two | Not specified | No information |
| North East 1 | Partner allowed (12 and 20 weeks)<sup>†</sup> | Unaccompanied | No information | One | Not specified | No information |
| North East 2 | Partners allowed | No details | No information | One | Not specified | No information |

\* Phone call offered if clinical concerns
¶ Video offered of a small section at the end of scan
† Phone call offered (end of scan)

### Details of themes

Overall, six themes emerged from the final synthesis of the data sets. An overview of the documents and interviews that generated data for each theme is presented in Table 3.

**Table 3:**
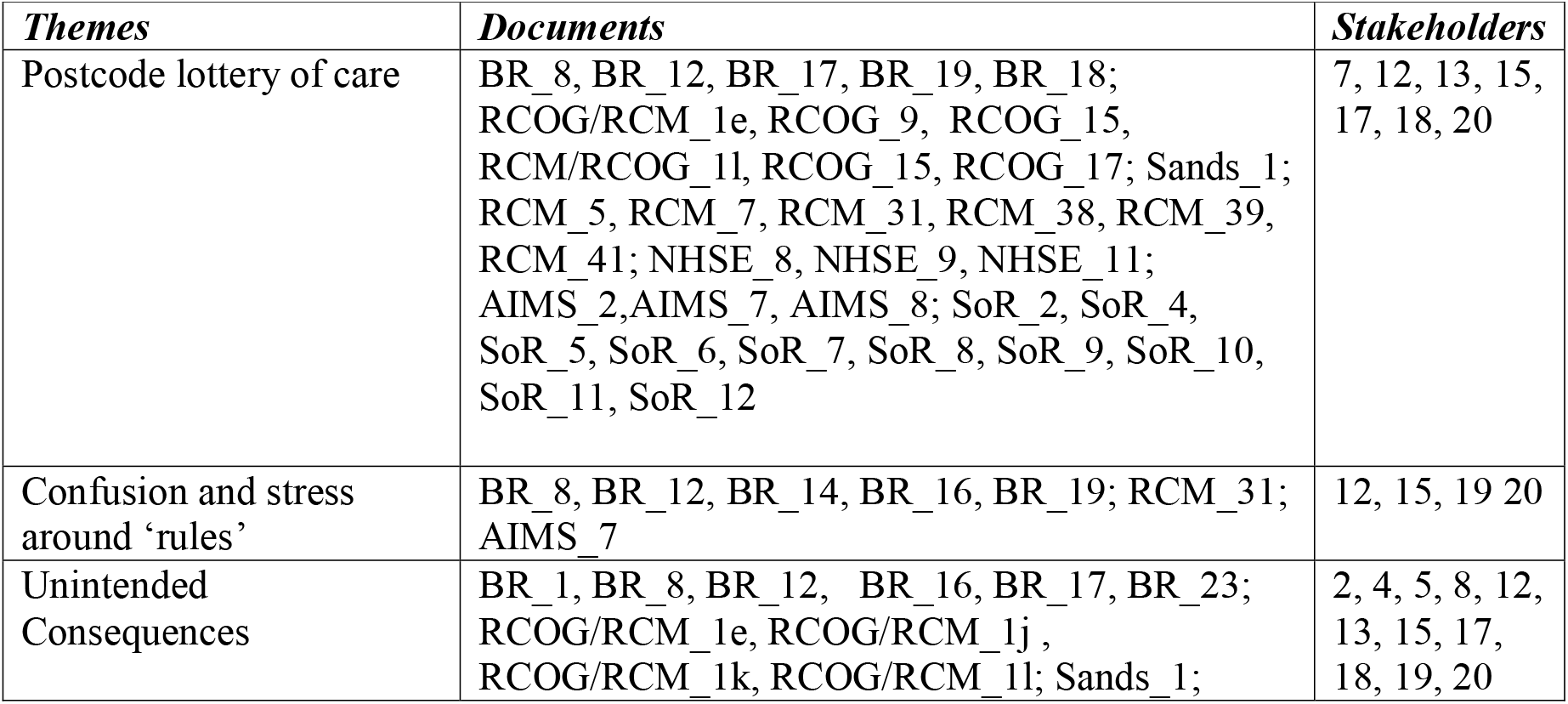
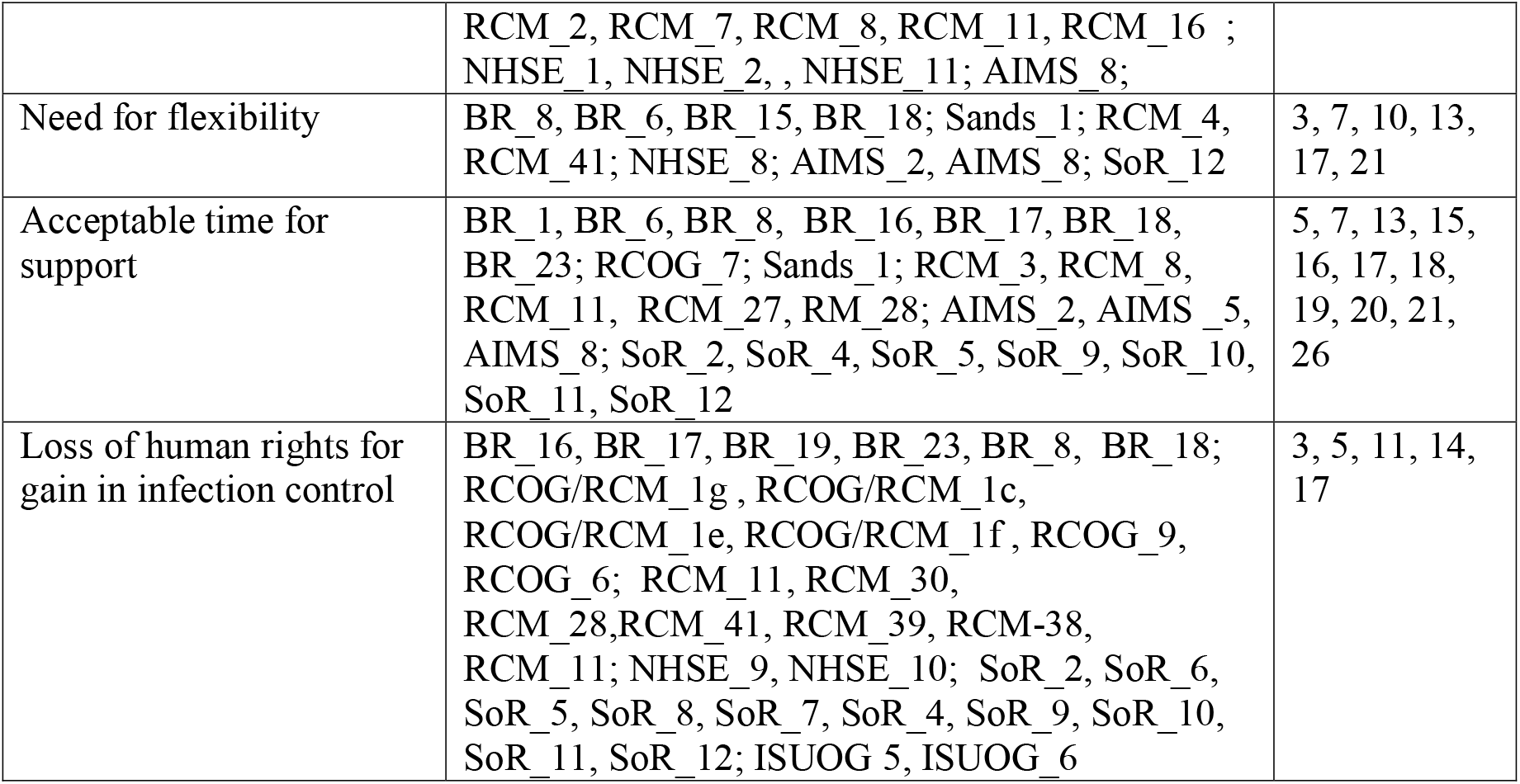
Documentary and interview data sources for the six key themes.

#### Postcode lottery of care

The notion of a postcode lottery of maternity service provision gained traction in the media over the summer of 2020 (16). Concerns were reflected in national documents from almost all included organisations, the stakeholder interviews, and reflected in Trust level responses. Variation was justified by some organisations as a reaction to local need, for example: *‘restrictions on other visitors should follow hospital policy and national guidance*’ (RCM_7) and ‘a*ll staff should work to the same local policy, to provide a consistent service to women’* (SoR_5). The caveat to most guidance was that any policies needed to be re-addressed in the event of local spikes in COVID-19 cases or following local risk assessment, to ensure a ‘*consistent service to women’* (SoR_2):

> But it’s guidance, and every hospital will make its own decisions and to a certain extent, they will have to because the physicality, the layout, the facilities that they have in hospitals will differ. You know, there’s not much room in the waiting area if the corridors are very narrow [so] that the people can’t have a two-metre distance. (Stakeholder 20, Miscarriage Association)

Some Trusts were identified as having ‘*gone out of their way to ensure their services remain family-centred’* (BR_14), whereas in others, partners were unable to attend any antenatal appointments or scans (BR_16). NHS England released guidance in September 2020 intended to assist trusts to reintroduce access for partners and visitors (NHSE_8. Some organisational responses claimed that this led to ‘*some Trusts starting to backtrack and reduce restrictions in maternity services’* (BR_17), while other Trusts continued to impose restrictions (BR_18) and often without a clear rationale for these variations:

> And that’s the problem. You… because down the street, down the road, you could get a very warm, empathetic ultrasonographer who says, of course, yeah, I realize how difficult is. You know, it doesn’t take much, and we just have lost it because people are stressed and there’s lots of reasons for it, but it’s just not good enough. (Stakeholder 18, Independent Midwifery Advisor)

#### Confusion and stress around ‘rules’

Concern over a lack of clarity in decision making and changes in policies around companionship were highlighted. For instance, letters from Birthrights to maternity leads (e.g. BR_12, BR_16, BR_19) repeatedly emphasised the need for clear reasons, evidence, and justification as to why decisions were being made. Concerns included that Trusts *‘acted too quickly to withdraw services’* and *‘decision-making has not always been proportionate or transparent’* (BR_18). While most Trusts made some reference to infection risk as the rationale for the restrictions, a sizeable minority (9/25) did not. Many Trusts offered no rationale as to why partners could attend some appointments but not others, e.g.:

> One birth partner may attend for the 20-week anomaly scan only. Women to attend all other scan appointments unaccompanied (South West 2).

Birthrights and some stakeholders highlighted a failure to communicate local restrictions in a timely manner, compounded by rules changing rapidly and difficulties in communicating these changes widely and consistently to large numbers of healthcare professionals. This confusion about the rules at local level was also compound by changeovers of staff and communication between different teams in some areas – *‘there’s been no consultation with sonographers in terms of risk assessments* […] *or changes in practice’* (Stakeholder 26, The Society of Radiographers), and services being provided by staff from outside the maternity team.

While some stakeholders noted that individual Trusts responded to this confusion by using a range of public communications, data from the 25 maternity trusts found that less than half (10/25) had a consistent message about companionship across different channels. Parent frustration reported by stakeholders also related to how the rules for birth companions and visiting seemed at odds with the social distancing rules outside of the hospital context:

> In the middle of lockdown, it was ‘we don’t like it, but we know you’re keeping us safe’. Now ‘it’s we don’t like it and I don’t see how you’re keeping us more safe doing this because I can meet my partner in the pub, but he can’t come to my scan. I can do this, but I can’t do that’. Yes. So, it’s more of an angry mood now. (Stakeholder 12, Maternity Voices Partnership)

In some of the documents by Birthrights (e.g. BR_14; BR_16) and RCM (i.e. RCM_31) they called for the harm caused by restricted access by birth partners to be properly and transparently considered within the decision-making processes:

> We would be grateful if you could publish or send us the risk assessment that quantifies the increased risk of spreading COVID within the unit (despite PPE and other mitigating factors, and the fact that most partners are from the same household) caused by relaxing restrictions, and weighs this against the known harms to pregnant women, birthing people and their families from keeping the current restrictions in place. (BR_16)

#### Unintended Consequences

Organisations including RCM, RCOG, NHS England and Birthrights highlighted that having trusted companions throughout labour and birth is linked to improved outcomes, and a lack of companionship was associated with increased need for pharmacological or other interventions (AIMS_8, BR_23). This included perceived impacts on the labour process due to, for example, [increased] *‘demand for epidurals’* (RCM_8). Alongside the obvious fear and anxiety of infection, organisations and stakeholders highlighted concerns about women feeling ‘*petrified’* (Stakeholder 15, Professor of Midwifery) or not accessing maternity care, ‘*due to the women’s preferred birth partner not being allowed to accompany her*’ (RCM_2). There were also concerns of partners feeling *‘unsupported and uncared for’* (Stakeholder 20, Miscarriage Association) due to being unable to be with their partner when they heard bad news (during ultrasound) or missing the birth due to *‘being told that they should wait in the car park or something’* (Stakeholder 7, Birthrights). All but four Trust websites contained messages of empathy regarding the restrictions, sometimes alongside expressions of regret and/or justifications for their necessity: ‘*We understand the restrictions we have had in place over recent months have been particularly hard for pregnant women and their families*’ (North West 1). Concerns were also expressed by stakeholders and within documents by Sands and Birthrights, towards women who had experienced prior baby loss or for women for may receive bad news alone during the scan:

> Women are being asked to attend scans alone, with many sharing frustrations that they cannot video link to their partners. These very vulnerable women tell us they are concerned about having to attend stressful antenatal appointments and scans alone. While units are being encouraged to consider facilitating women to take a video clip at the end of an appointment, this is reliant on local policies. (Sands_1)

Companionship was noted to have practical as well as emotional implications for women. Some of the documents claimed that the absence of companions meant that women required more support from maternity care professionals (e.g. RCOG/RCM_1j; NHSE_1) creating additional stress for over stretched services (RCOG/RCM_1l), and additional potential exposure to COVID infection. Additional workload pressures were also created when staff had to police non-compliant behaviours:

> There have been frustrations. For example, partners now come on to postnatal ward for a fixed time period. And staff have reported things like literally having to find partners from behind curtains. “You need to go now”. … So, a lot of stress. Because [having to] explain to people why they can’t have what they have or what they want. (Stakeholder 12, Maternity Voices Partnership)

Some practitioner respondents reported that they or their colleagues experienced moral distress when social distancing rules prohibited physical contact with women who were alone, or receiving bad news:

> All of a sudden it was just women on their own for us. And that was really, really stressful for the women and the staff. And a lot of a lot of my job is giving bad news. And to give that to women that are on their own with no support; you can’t touch them. You can’t hug them. And so that for us is really, really challenging. I think that was probably the most challenging thing. (Stakeholder 24, Midwife Sonographer)

#### Need for flexibility

Concerns discussed within the documents (e.g. BR_8; BR_18; RCM_4) and raised by stakeholders related to the blanket adoption of visitation rules across whole hospitals. Some commented that pregnant women and childbearing people were a *‘separate population with separate needs’* (Stakeholder 7, Birthrights), arguing that visiting rules adopted in other areas of healthcare should not apply to a perinatal population:

> So, you know, several heads of midwifery were saying to me, I want to do this, but they won’t let me because they made a decision about what the visiting will look like in this hospital. And they do not see maternity as an exception. And, you know, it is an interesting reflection, isn’t it, that maternity has always been a service that has seen itself as an exception to the health care service in which it sits. (Stakeholder 17, NHS England and NHS improvement)

Responses from organisations including AIMS, Birthrights and the RCM argued how there needed to be consideration of women’s unique situations. Restrictions on companionship were considered to have a disproportionate impact on those who were facing disadvantages including those for whom English is a second language, women with mental health problems, cognitive impairments, refugee and asylum seeking women (AIMS_8, RCM_4, BR_8, BR_18). Only one Trust included a statement on their website about offering personalised (flexible/individualised) care for all women, that might not be in line with COVID policies. Five others said they offered this on a case-by-case basis (often expressed as ‘exceptional circumstances’). One of these referred to Black, Asian, and Minority Ethnic communities along with concerns about greater COVID risks, and three referred to ‘allowing’ women to bring a companion if they ‘needed assistance’. In two Trusts this was explicitly linked to women with disabilities:

> Partners and family members will not be allowed to enter the building unless you need support from a carer/relative (for example, if you have a disability and need support) (North West 2). Birthrights stipulated how ‘protected characteristics’ under the Equality Act 2010 (e.g. physical disability or mental health condition) meant that maternity Trusts were obliged to make reasonable adjustments (e.g. BR_15; BR_18). NHS England emphasised the need for ‘essential visitors’ (seen as different to ‘normal visitors’) for those with specific communication or care needs (NHSE_8). AIMS also stressed that on some occasions, on a ‘*case by case basi*s’, such as a disability, *‘a second birth partner may be critical to women’s mental wellbeing or other needs’* (AIMS_2).

The lack of flexibility for highly sensitive events such as for those who experienced trauma or loss were also highlighted by Birthrights (e.g. BR_18) and RCM (e.g. RCM_41). While some incidents of positive practice were identified, stakeholders also shared stories of parents whose baby had died in utero being unable to take photographs or spend time with their deceased child:

> We had a lot of stories from parents who hadn’t been allowed to take photographs, haven’t had time to sit and hold their baby. And I think all of those were linked to both a lack of space, to lack of bereavement space, but also a lack of staff understanding of how to adapt bereavement care standards. We also saw in this group a lot of problems around not having the partner with them. (Stakeholder 10, Sands)

#### ‘Acceptable’ time for support

Access to and timing of support was an issue at key stages during the perinatal journey, and notably during antenatal ultrasound appointments and during early onset of labour.

A key area of contention related to women having to attend ultrasound scans unaccompanied: a situation described by one of the stakeholders as *‘ludicrous’* (Stakeholder 7. Birthrights). RCOG guidance recommended that *‘patients should be asked to attend alone if possible or with a maximum of one partner/visitor’* (RCOG_5), whereas a RCM document stated ‘*partners should attend scans unless rooms are too small to socially distance: partners may attend scans virtually’* (RCM_28). However, in contrast to this permission for virtual contact, a joint statement by SoR, RCOG, RCM and the British Medical Ultrasound Society stated that devices required for remote contact by partners via video/phones are a vehicle for transmission (due to surface contamination), and that recordings would impact on scan time, sonographer concentration and potential detection of fetal abnormalities – although it was acceptable for the mother (if in line with local policies) to “*save a short 10–30 second cine clip of the fetus at the end of selected examinations”* (SoR_11).

Trust data revealed that while most permitted partners at one or both standard ultrasound appointments (12 and 20 weeks), seven (∼30%) did not. Four Trusts mentioned video or other means of ‘virtual’ companionship, but usually to specify that videos of scans were not permitted. Only one Trust referred to women being able to phone a partner for support if the sonographer were to find “*important clinical information that your partner needs to be aware of*” (Greater London 2). While many stakeholders were critical of the ultrasound restrictions on companionship, one respondent argued the need to highlight that antenatal ultrasound scanning continued, even when *‘other screening programmes went into hibernation’* (Stakeholder 26, The Society of Radiographers). Some defended restrictions on companionship during scanning, noting that scans often have to take place in areas *‘like a broom cupboard in a very small poorly ventilated space’* (Stakeholder 17, NHS England and NHS improvement) coupled with the restricted time to undertake the examination and sterilising the room and equipment after each appointment. One reported that there had been a *‘downgrading’* of the importance of scans as a medical examination that required focused concentration in challenging situations, during vociferous debate about partner attendance (Stakeholder 26, The Society of Radiographers). However, while sonographers may have faced increased risks due to screening large numbers of women, the specific rationale for not allowing videos as an alternative was challenged:

> You can argue the toss as to whether some of the justifications for not allowing that were real or weren’t real. You know, is there really a risk of infection if you pick up your phone? Really? Maybe some anxiety for sonographers or whoever’s doing the scan. You know, you don’t really want the phone on with a video while you’re doing the scan because who knows, they might use it in some kind of litigation. Who knows? But whatever it was, it really didn’t help. (Stakeholder 20, Miscarriage Association)

A further area of contention concerned companionship during labour and birth. While organisations such as Birthrights argued for companionship throughout, less than half (9/25) the Trusts referred to partners attending ‘*throughout*’ or ‘*for the duration*’. Three Trusts referred to companionship being permissible only when the woman was in ‘*established*’ or ‘*active*’ labour (with no details as to how this would be established); and 13 Trusts did not specify the relevant phase of labour. RCM guidance advised that women would not be able to have partners present during inductions that took place in a bay or ward (RCM_27). Only six Trusts (25%) indicated that partners could be present during induction of labour, and four allowed partners, but with limitations (either restricted to daytime or if the woman needed additional support). About half (12) provided no information, and three explicitly stated that partners were not allowed:

> If you are attending for induction of labour please attend alone, your birth partner will join you once you are transferred to the Delivery Suite. (Yorkshire and Humber 1)

Birthrights and AIMS (i.e. BR_8; AIMS_5; BR_18; BR_23) also raised concerns about cervical dilatation as the only acceptable indicator of active labour, meaning that some women who may not have wanted (or needed) a vaginal examination felt pressured to accept the procedure if they wanted their birth companion to be granted access to the labour ward.

There were examples of innovation to try to support companionship for women and access for their partners. Some Trusts initiated or extended the provision of labour induction in community settings or in private hospital rooms (rather than multi-occupancy early labour wards) to prevent separation of women from birth partners (RCM_8; RCM_27). Some stakeholders also referred to more flexible approaches to induction such as partners being able to *‘come and settle them* [women] *in’* and to use *‘Facetime to be with their partner all the time’* (Stakeholder 21, Midwife with a national role).

#### Loss of human rights for gain in infection control

There was some evidence from stakeholders that hospital decision makers in some settings believed that companionship during the maternity episode should not be prioritised over other areas where attendance of close family members would usually be seen as a critical human need and right: especially when someone was dying in hospital:

> So, it was interesting and when I would speak to the head of midwifery, sometimes it felt like they were saying, you know, well, everyone’s got to make….sacrifices. And there are people dying alone in hospital. There are people suffering terribly alone in hospital, unable to have visitors…. [while] there were women saying, you know, it’s my right to have a companion it’s your job to provide care for me. So, it felt at times like each group with their own concerns was unable to think about or found it difficult to take on board the concerns of the other group. (Stakeholder 17, NHS England and NHS improvement)

The underlying principle within most of the guidance reviewed was that ‘safety’, was primarily conceptualised as the prevention of transmission of infection, both to service-users and to staff. NHS England documents referred to minimising ‘*control risks working with your IPC* [Infection, Prevention & Control] *leads, whilst still allowing the maximum possible safe access’* (i.e. NHSE_9). The RCOG/RCM also noted the need to minimise the number of attendees (as ISUOG_5 quote above), but acknowledged that one person could be there for antenatal visits should a woman choose this:

> You will be asked to come alone to clinical appointments or keep the number of people with you to one (including midwifery visits in your home). This will include being asked not to bring your children with you to appointments. This is important to protect maternity staff, other women, and babies, and you and your family from the risk of infection. (RCOG/RCM_1g)

Birthrights was one of the key organisations to recommend that notions of safety might also include emotional and psychosocial risks of women being unattended (e.g. BR_17, BR_16, BR_19, BR_18) - ‘*The damage caused by ongoing restrictions needs to be weighed up against the requirements of infection control’* (BR_19). Several of their documents (e.g. BR_18; BR_19) claimed that routinely restricting birth companions was a violation of women’s (and partners) human rights. Despite this, stakeholders stated that, in practice, women’s human rights and choices around companionship did not feature as part of the decision-making processes:

> And I think we’ve spent the last, you know, however many years banging on about the fact we want to give women choice and rights and sharing that discourse and encouraging women to become empowered. And then COVID comes along and we just say, no, no, we’re not doing that (Stakeholder 17, NHS England and NHS improvement)

The RCM (RCM_41) stated that their ‘*greatest concern’* was ‘*safety being sacrificed in favour of popularity’* which seemed to imply that companionship should not outweigh the need to prevent infection of its *‘members’* and of ‘*women and families’*. SoR also highlighted that its guidance had *‘risk assessments’* at its core (SoR6). The guidance did not preclude ‘people being accompanied’, but that it ‘*must only happen if the safety of the patient and sonographer is not compromised*’ (SoR_8). However, others argued that day to day decision making was based around a belief about safety that was limited: ‘*because it’s not just about the physical self, it’s about psychological self*’(Stakeholder 14 Senior Midwife Leader):

> But I think safety generally is an interesting thing because….You know, so many different things affect safety don’t they, so something like being able to have your partner with you might not be seen as a primary thing affecting safety in comparison with protecting against COVID, but actually if it impacts on someone’s mental health in either the partner or the mother, that does have an effect on safety (Stakeholder 5, National Maternity Voices Partnership)

## DISCUSSION

In this paper we have drawn on guidance from national statutory and service-user organisations, key stakeholders, and public facing Trust-level data to consider the organisational issues associated with companionship in maternity care during the COVID-19 pandemic. The value of companionship during labour and birth for women and birthing people is widely recognised, in terms of clinical benefits, and short and longer term psychosocial impacts (1, 9). As evidenced within this paper, during the COVID-19 pandemic, at the policy and organisational level, assumptions and norms about companionship, accompaniment and visiting during facility-based health care provision have faced profound challenge. Others have noted the variance in maternity organisation response during the pandemic(17). Some variation can probably be explained by changing national knowledge about the prevalence and impacts of COVID-19, and by different levels of exposure to COVID-19 infection. However, our data suggest that this was not the case where blanket policies were applied with minimal individual flexibility, or where there was unjustified variation in visiting and companionship rules, coupled with poor and inconsistent communication.

We found particular concern about lack of access to companionship in two distinct areas. First women being unable to have any communication (actual or virtual) with partners or other family members at ultrasound scan; and denial of intrapartum companionship until labour was ‘established’. In relation to the former case, there is some evidence that, beyond the emotional and psychological benefits for the mother, when fathers and partners are present for antenatal ultrasound scan, there are significant effects on their identification with the fetus (as their future child) and their empathic relating with the mother(18, 19). This implies that being present for ultrasound scans has important public health and relationship benefits for them, and their partner and baby. In the latter case, in some Trusts, ensuring that labour had progressed sufficiently was perceived by some stakeholders to be associated with coercive and invasive practices, such as regular vaginal examinations when women may otherwise not have needed or wanted such examinations. General uncertainty over organisational companionship permissions may also be reflected in anecdotal rises in women choosing to freebirth(20, 21), and the issuing of associated RCM guidance to ensure appropriate professional responses(22). Trust policies that restricted companionship until labour was established (or until birth was imminent) seemed to be built on an assumption that companionship was only really needed when labour was very intense, and/or when the birth was imminent so that the birth companion (as the co-parent) could be ‘permitted’ to witness to the birth of their baby. In contrast, other Trusts seemed to recognise, at the organisational level, that active and engaged birth companionship throughout labour (from the early stages of spontaneous labour, or from the time of labour induction through to the birth) is a mechanism for clinical, psychological and emotional safety for the mother, partner and child, both in the short term, and, critically, in the longer term, when the threat of COVID-19 infection is long over (11, 23).

The pandemic brings into sharp focus the fundamental and underpinning ethical dilemma between social actions that ensure the greatest benefit for the population as a whole, and the individual human rights of each person within that population(24). Resolving this potential conflict of ethical imperatives depends on an open and informed debate about rights and consequences. In terms of maternity care, this requires a sophisticated understanding of what ‘companionship’ means, over the whole life course, and for mother, baby, and family. It also requires attention to the potential moral distress of maternity care staff (and health care staff in general, including ultrasonographers) who are faced with the stress of having to balance these two imperatives with real people, in intensely emotional real time, repeatedly day in and day out, and at times with insufficient PPE equipment available; at a time when they, too could be pregnant women at risk of exposure to infection, or parents/carers fearful of infecting family members(25-28). The study by Nguyen and colleagues identified that that infection risks for acquiring infection were high for staff compared to women, and particularly among front-line health-care workers(29).

This is the first study to bring together national policy and stakeholder views with Trust based public facing data to understand how maternity care has been organised in England during COVID-19. Although we cannot be sure we captured every single relevant document produced over the period of our data collection, triangulation across data sources and across multiple researchers enabled rich insights into how and why variations occurred, and the perceived impacts. The pragmatic restriction of the Trust level data collection to only 25 Trusts (10% of maternity care providers in the UK), and the restriction to maternity-specific documents and guidance may be seen as a limitation, the organisations that were included were selected purposively to reflect a wide range of relevant characteristics. Since this paper is focused on policy and organisational responses to the pandemic, service-users, birth companions and healthcare professionals were not included. In addition, our analysis did not include findings related to postnatal care, or care in neonatal units. These areas, and the unintended (positive and negative) short- and longer-term consequences of different interpretations of the value of companionship when balanced against infection control, are critical areas for examination during the on-going COVID-19 crisis. Future outputs from the ASPIRE project will address these gaps.

## CONCLUSION

This paper presents insights from the ASPIRE COVID-19 UK study to understand how companionship in maternity care was operationalised at the organisational level in antenatal and intrapartum care during COVID-19. Our findings illustrate variations in policy at national and local level, coupled with poor and inconsistent communication of how the restrictions changed in some sites, and a lack of clarity in the decision-making processes. The evidence highlighted a lack of flexibility in responding to women with more complex needs in some cases, the negative and positive unintended consequences of companionship restrictions, and the challenges of conceptualising and balancing infection risk and emotional and psychological distress. However, there was evidence that creative solutions were possible, since, despite significant pressures, some Trusts appeared to continue to provide full companionship.

Overall, these concerns illustrate something much more fundamental than merely barriers to hospital ‘visiting. While the NHS England Better Births policy agenda highlights the need for safety and personalisation within maternity care, these findings suggest that, over the time period captured by this study, personalisation (and emotional and psychological safety) became sacrificed in some (but not all) situations to the overriding imperative to minimise infection spread with high emotional and psychological costs. Further research should capture the views and experiences of healthcare professionals and service-users, and clinical outcome data from different settings. There is an urgent need to determine how to balance risks and benefits sensitively and flexibly and to create optimum outcomes for women, pregnant people, fathers, parents, infants, families and staff, during the current and future crisis situations.

## Data Availability

Trust level data is included in the paper. Details of all documents analysed is provided/with all information freely available. All relevant interview data concerning companionship will be openly available from UCLanData and a doi provided on paper acceptance.

## Author contributions

SD with input from the ASPIRE-COVID19 Collaborative Group designed the study. RN, GT, SD and NC interviewed the stakeholders; MCB, GM and JC were involved in data extraction for the documentary analysis; SH, AS and NC collected and analysed Trust level data. GT, MC and RN were involved in developing themes from the interview and documentary data, and GT synthesised the Trust level data into the data set. Final themes were agreed with all authors. All authors and the ASPIRE-COVID19 Collaborative Group contributed to writing and reviewing the manuscript.

