## Supplementary File 1 for "Companionship for women using English maternity services during COVID-19: National and organisational perspectives"

### Interview schedule

**Opening question**

In your view, what are the most important issues for maternity care provision that have come out of COVID-19?

Probe: *Are these views based on any particularly memorable experiences?*

**Changes/adaptions to service delivery**

How have [i.e. maternity, midwifery, obstetric, neonatal, ambulance delivery, staffing, any other?] services been adapted during COVID-19 pandemic?

Prompts:	*How significant or minor have these changes been?*

*Can you describe some of the primary Covid-19 interventions?*

*Have these adaptions varied by place and/or time? If so, what do you think explains this variation?*

**Decision making processes**

From your perspective, who makes decisions about the changes?

Prompts:	*How are these decisions made?*

*Were decisions made at a multidisciplinary level or separately?*

*How did the different agencies – work together - who had overall responsibility?
Who has been consulted about the changes – are all those impacted consulted (e.g. at UK level were MVPs consulted and/or for neonatal were the local PAG group consulted?)*

*If there is resistance, how is that dealt with?*

How have safety principles (for service-users and staff) informed decision-making/practice changes?

How have personalised care principles informed decision-making/practice changes?

Prompts:	*What personalised care aspects have been considered?*

To what extent has the needs and care of BAME/vulnerable populations been considered in the decision-making processes?

What other factors have influenced decision-making/changes in care provision?

**Communication and implementation**

How have the recommended practices and guidance been developed?

Prompts:	*How effective have these methods been?*

*How was conflict in what to include resolved?*

How have they been communicated to practitioners/front line staff?

Prompts:	*How effective have these methods been?*

*What more is needed?*

How is service delivery being monitored over this period?

Prompts:	*What feedback loops are in place*?

*How is this information used?*

**Impact**

What has been the (positive and negative) impact of the changes/adaptions on:

- Staff
- Parents and families (siblings/grandparents)
- Service delivery
- Data collection and sharing (i.e. Maternity Services Dashboard and Better Births Minimum Data set)

In which data do you think the effects of the pandemic and the adaptations made to service provision can be seen?  Can we have access to the data?

**Barriers and facilitators**

What has worked well in providing safe and personalised maternity care during the pandemic?

Prompts:	*Do any particular Trusts/services/lobby groups and etc stand out for you? Why is this?*

What have been the challenges or barriers in providing safe maternity care?

What have been the challenges or barriers in providing personalised maternity care?

**Recommendations and Sustainability**

What else is needed to ensure safe and personalised maternity standards and principles are upheld in:

- future routine maternity care
- a future crisis/pandemic

If there was one thing that you would change about how maternity care was managed what that would be?

What innovations been implemented that could be opportunities for how maternity care could be organized in the future?

Prompts:	*Can you identify where these innovations and opportunities are happening at the moment?*

How do you think services should prepare for future occurrences/pandemics?

Is there a specific Trust/Health Board you think we should include in our case studies?

Prompts:	*If so, can you tell us why you think this is a particularly relevant site?*

Are there other things, that have not been mentioned that you would like to share?

Is there anyone else that you think we should talk to?
